# Electroencephalographic features of chronic subjective tinnitus: A scoping review

**DOI:** 10.1101/2025.03.24.25324557

**Authors:** Lynton Graetz, Mitchell Goldsworthy, Kenneth Pope, Sabrina Sghirripa, Tharin Sayed, Rebekah O’Loughlin, Giriraj Singh Shekhawat

## Abstract

**Objective:** The goal of this scoping review is to review the scope of features from previous resting-state electroencephalography (EEG) research that have the potential to be objective measures of chronic subjective tinnitus and presence.

**Methods:** Using keywords related to resting-state EEG and tinnitus we retrieved studies from Ovid, PubMed, and CINAHL. Studies were included if they met the following criteria: studies utilized resting-state EEG to assess chronic subjective tinnitus symptoms or compare those with tinnitus to control subjects.

**Results:** We identified and reviewed 81 comparison studies that used resting-state EEG. Spectral power source and electrode analysis were the most common results reported in studies that compared those with chronic subjective tinnitus to controls. Connectivity and network analysis were the third most reported analysis type.

**Conclusions:** Electroencephalography (EEG) has potential to be utilised as an objective measure of chronic subjective tinnitus, independent of subject reports. While no single definitive EEG marker for chronic subjective tinnitus was identified, emerging evidence from comparison studies indicates a strong impact of chronic subjective tinnitus on network measures and connectivity related features. We propose a pathway to establishing objective measures from EEG recordings. Combining spectral band power, network and connectivity as features and validating them with machine learning classification on large, diverse populations, is a proposed pathway to establish objective measures.

**Significance:** Idiopathic chronic subjective tinnitus affects up to a 20% of the population, particularly in the elderly, is associated with hearing loss, and lacks widely accepted treatments. Assessing tinnitus, and symptom changes are challenging due to the lack of objective measures of idiopathic chronic subjective tinnitus. This review focuses on the use of resting-state EEG research to identify candidates that may be developed for use in research and by clinicians.

## 1. Introduction

Tinnitus is a condition in which a phantom sound is heard with the absence of any external source (Baguley et al., 2013). Idiopathic chronic subjective tinnitus (defined as no identifiable cause, and persisting for longer than 3 months) can reduce quality of life (Langguth, 2011), and is highly prevalent, affecting up to 25% of the Australian population, with 7% of working Australians experiencing constant symptoms (Lewkowski et al., 2022), and 10% to 15% prevalence reported worldwide (Baguley et al., 2013). Some forms of tinnitus have an identifiable cause; however, most cases are idiopathic (of unknown cause) and subjective, heard only by the individual. The auditory system generates a phantom perception of sound and individuals report a wide variation of pitch, volume, and tone (Hazell & Jastreboff, 1990; Langguth et al., 2007). Often associated with hearing loss or sound exposure, models suggest chronic subjective tinnitus is a compensatory mechanism for nerve fibre damage and a result of deafferentation of auditory input (Paul et al., 2017).

There is currently no universally effective treatment to relieve the symptoms of idiopathic chronic subjective tinnitus, and no objective measure of chronic subjective tinnitus. Current methods of tinnitus assessment rely on self-report using questionnaires such as the Tinnitus Handicap Inventory (THI) (Newman et al., 1996) and the Tinnitus Functional Index (TFI) (Meikle et al., 2012), subjective audiological measures of pitch matching, loudness matching and minimal masking levels (Cima et al., 2019), and visual analogue scales of loudness and annoyance. These subjective measures are useful, but the development of cost-effective objective markers of chronic subjective tinnitus would aid in tracking changes in symptoms, identifying susceptibility, stratifying subtypes and personalised treatment for tinnitus (Husain & Khan, 2023), and assisting medico legal purposes (Fabrizio-Stover et al., 2024; R. Jackson et al., 2019).

Differences in neural activity among individuals with chronic subjective tinnitus can be objectively observed. Among measurement methods, resting-state electroencephalography (EEG) stands out for being non-invasive, relatively cost-effective, accessible, and requiring no participant input. EEG at the sensor level measures neural signals by recording the electrical post-synaptic currents in the brain’s cortex through the potentials generated at external scalp sensors. Source level analysis (Grech et al., 2008) enables the estimation of deeper brain sources and subcortical structures from surface EEG recordings, offering valuable insights into the neural origins of tinnitus. This method has been widely adopted in tinnitus research to investigate activity in regions such as the auditory cortex and limbic system. However, source analysis remains a topic of debate due to challenges such as variability in methodological approaches and the inherent limitations of EEG in precisely localizing deep brain activity. These factors underscore the need for cautious interpretation.

The search for objective measures of chronic subjective tinnitus has led to diverse approaches. A recent systematic review on objective measures of tinnitus by Jackson et al. (2019) identified 21 articles assessing objective measures. The methods reviewed included measures derived from blood samples, radiological scans, balance assessments, and electrophysiological tests. Fifteen of the 21 studies employed EEG-based measures: five evaluated quantitative resting-state EEG metrics, while the remaining ten employed evoked or event-related EEG measures. The conclusions of this review echoed a previous scoping review on hearing aids (Jacquemin et al., 2021), indicating that no reliable or reproducible objective measure for chronic subjective tinnitus has been established (McCormack et al., 2016). Jackson et al (2019) recommended that further research is necessary, potentially incorporating tools like fMRI or EEG, to complement self-report questionnaires in tinnitus assessment Magnetoencephalography (MEG) is a similar method resulting in less noise compared to EEG. A recent review of tinnitus related MEG findings (Reisinger et al., 2023) found initial studies reported increased resting-state alpha and delta band power, suggesting hyperexcitability and reduced alpha variability, although these findings were tempered by inconsistent results in subsequent studies. Increased functional connectivity between auditory and non-auditory networks and in alpha and beta bands was also noted, suggesting auditory regions alone are not sufficient for understanding tinnitus (Reisinger et al., 2023).

Resting-state EEG has been extensively utilized in tinnitus research to evaluate the efficacy of various treatments, including acoustic, electrical, and magnetic stimulation, as well as psychological interventions and neurofeedback. It also aids in exploring neurophysiological differences in individuals with chronic subjective tinnitus (De Ridder & Vanneste, 2021; Elgoyhen et al., 2015; Fabrizio-Stover et al., 2024; Güntensperger et al., 2017; Husain & Khan, 2023; Lan et al., 2021; Lobarinas et al., 2008; Yasoda-Mohan & Vanneste, 2024). However, no recent reviews have summarised the findings from resting-state EEG measures in tinnitus research. This scoping review aims to examine the current literature to identify potential neural markers of chronic subjective tinnitus and chart a course toward clinically applicable objective measures.

Our review was structured according to the methodology of Arksey & O’Malley (2005). The objectives were to:

- Compile research on EEG markers from resting-state comparison.
- Identify resting-state EEG derived features observed in those with chronic subjective tinnitus compared to controls.
- Suggest future research directions to develop reliable clinical EEG tools for diagnosing, monitoring, and subtyping tinnitus.

## 2. Methods

### 2.1. Inclusion exclusion criteria

Using a systematic approach to addressing the question, without excluding studies based on quality (Mays et al., 2001; Munn et al., 2018), we followed a five stage framework. Stage 1: Identifying the research question; 2: Identifying relevant studies; 3: Study selection, 4: Charting the data, 5: Collating, summarizing and reporting the results (Arksey & O’Malley, 2005).

The PRISMA-ScR checklist was used to guide reporting (Page et al., 2020) and Covidence (Goulas, 2022) was used as a tool by the reviewers to maintain the workflow and extract data using standard forms. The inclusion and exclusion criteria are in Table 1. This review included all resting-state EEG-derived features, including frequency spectrum measures, functional and global connectivity measures, information theoretic measures and microstates, to comprehensively identify indicators of tinnitus and identify emerging themes.

**Table 1.** Article eligibility criteria.

| Inclusion | Exclusion |
| --- | --- |
| Subjective non-pulsatile tinnitus | Objective tinnitus (i.e. tinnitus with a known cause) |
| Chronic subjective tinnitus | Case studies |
| Resting-state EEG | Event-related potential (ERP) studies (i.e., not resting-state EEG) |
| Human studies | Non-human studies |
| Primary research studies | Reviews |
| Available online | Methods papers |
| Available in English | Magnetoencephalography (MEG) |
|  | Invasive recordings (i.e. electrocorticography or ECoG) |
|  | Intervention Studies |

### 2.2 Formulating the Research Question

The research question is: Can objective measures of tinnitus be identified using resting-state EEG features in individuals with tinnitus?

### 2.3 Identifying relevant studies

A research librarian assisted in the development of search strategies using relevant keywords and medical subject headings, focusing on EEG and tinnitus. Broad terms were used to capture as many studies as possible that used resting-state EEG in the evaluation of tinnitus, tinnitus symptoms, and change after treatments. No start date was set. The first study to fit the criteria was 2001 the end date of the search was 18 October 2024. For this scoping review, we selected PubMed, CINAHL, as primary databases for their extensive coverage of health-related literature, aligning with the review’s focus on tinnitus and EEG. Embase was chosen to compliment these databases with a broader range of interdisciplinary research.

### 2.4 Search terms

The search contained the following search terms; *tinnitus.mp. [mp=title, abstract, full text, caption text] and tinnitus.m_titl. and eeg.mp. [mp=title, abstract, full text, caption text] or electroencephalography.mp. [mp=title, abstract, full text, caption text]* on Ovid. The search terms were replicated for searches in the PubMed *(“tinnitus” AND (“eeg” OR “electroencepalography”))*.

*TI tinnitus AND TI ( eeg or electroencephalogram or electroencephalography ),* CINAHL <u>(Tinnitus AND (eeg or electroencephalography or electroencephalography)),</u> and Embase. *(((tinnitus[MeSH Terms]) AND (tinnitus)) AND (electroencephalography[MeSH Terms])) AND (electroencephalography) databases*.

We built the search terms using variations of the theme and related review articles.

### 2.5 Study selection

The results of the search and screening for eligibility criteria are shown in Figure 1, resulting in 81 included studies from 879 references.

**Figure 1.**
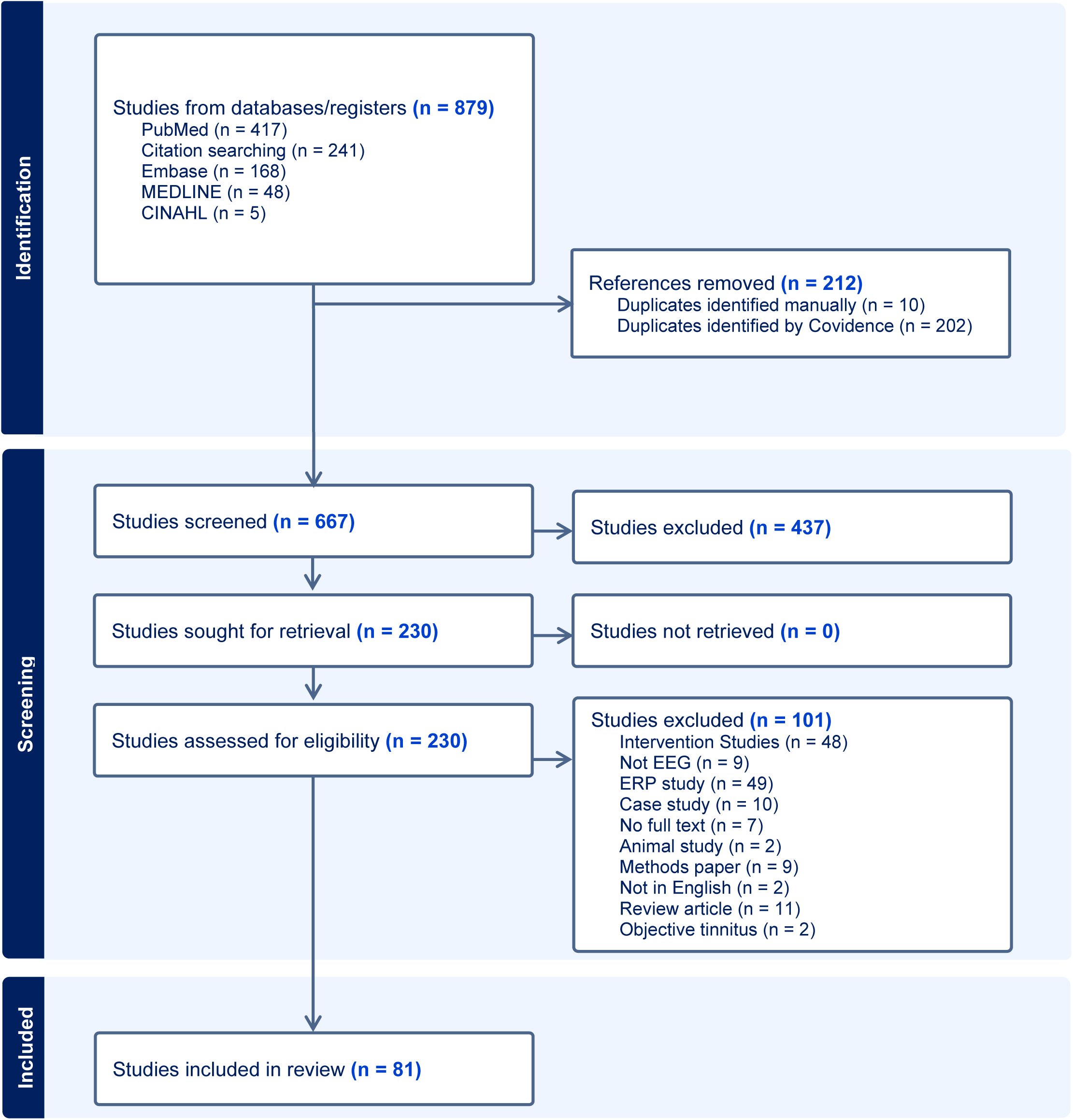
PRISMA Extraction diagram.

### 2.6 Data Extraction

We reviewed the 81 papers to capture the main EEG features of difference between groups using Covidence (Goulas, 2022) templates. The focus for this review was on EEG measures of chronic subjective tinnitus that were derived from resting-state EEG, and that made their comparison between participants with tinnitus and controls (between subject design). Some comparison studies were between related features or demographics such as EEG differences in gender, hyperacusis, hearing loss level, onset characteristics, sidedness, anxiety, distress and depression, coping style, loudness, frequency of tinnitus, and the ability to turn tinnitus on and off. These studies are included in Appendix 1 for reference but were not included in the results, comparison tables or figures. Unless indicated otherwise tinnitus refers to chronic subjective tinnitus.

## 3. Results

81 studies made group comparisons between people with tinnitus and controls using EEG feature analysis such as spectral band power at source and sensor level, sensor and source reconstructed functional connectivity, network analysis, microstates, and entropy (Figure 2).

**Figure 2.**
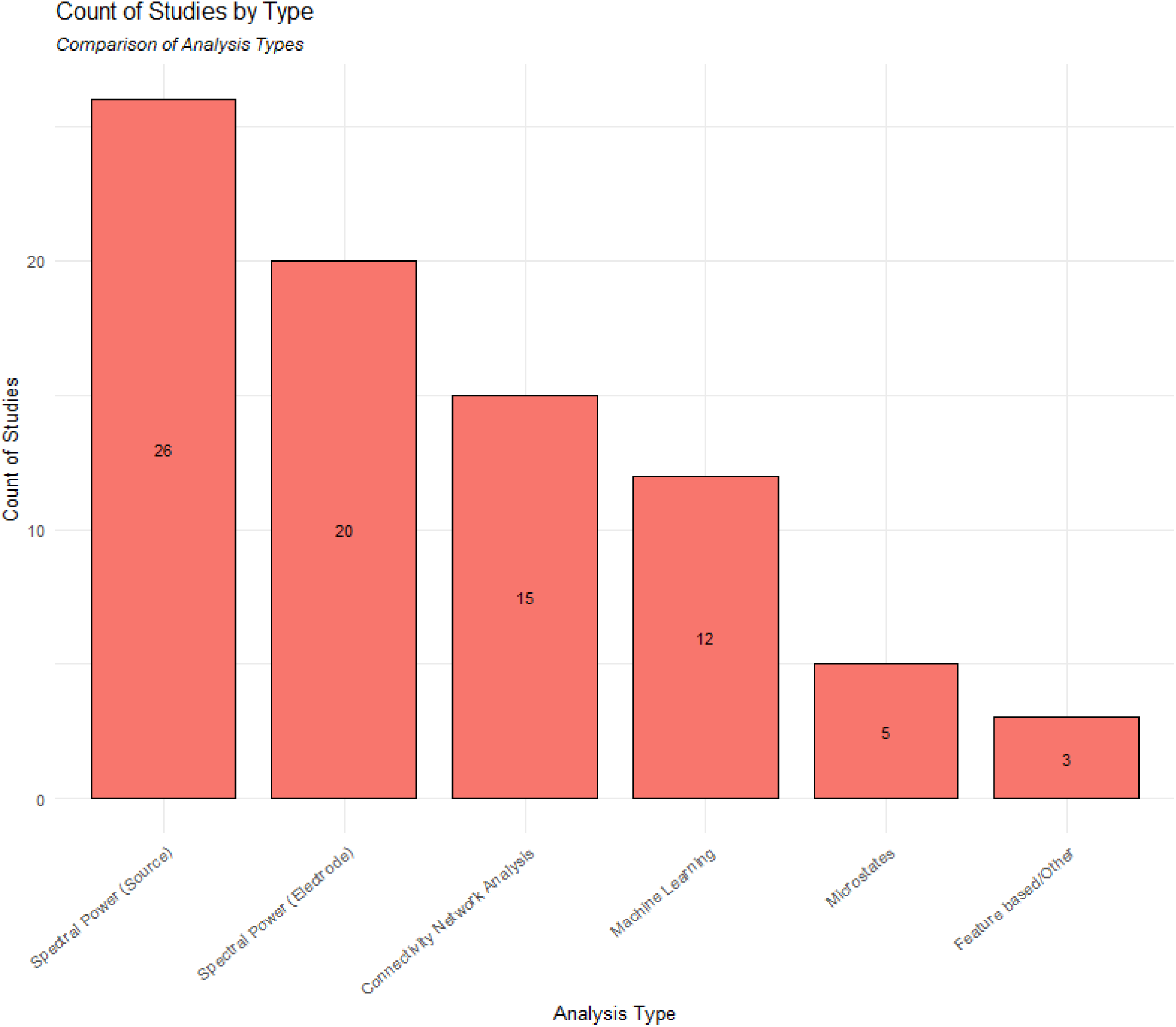
A histogram of studies using EEG to evaluate tinnitus. Several studies reported both band-power and connectivity results.

The summarized EEG spectral band power analysis of tinnitus compared to controls. Studies using EEG analysis comparing tinnitus to controls using connectivity and network measures are in Figure 4 and Table 2. Studies comparing Machine Learning techniques or using multiple features for tinnitus classification are in Table 3. Information theory studies are in 3.5 and microstate studies are in Table 4.

**Table 2.** denotes connectivity between: **General Terms:** TN tinnitus, C control, L left, R right, THI Tinnitus Handicap Inventory, **Auditory Pathways**: AC auditory cortex, IC inferior colliculus, STG superior temporal gyrus, TP temporal pole. **Frontal Areas**: FC frontal cortex, VLPFC, ventrolateral prefrontal cortex, FMC frontal medial cortex, dACC dorsal anterior cingulate cortex, pgACC pregenual anterior cortex, CCG central cingulate gyrus, M1 motor cortex, SMA supplementary motor area, IFG, inferior frontal gyrus. **Parietal and Sensory**: PFC parietal frontal cortex, SPG superior parietal gyrus, SOM somatosensory cortex, ITG inferior temporal gyrus, STG superior temporal gyrus, V1 primary visual cortex, V2 secondary visual cortex, IPS intraparietal sulcus. **Limbic System and Emotional Processing**: PCC posterior cingulate cortex, HIP hippocampus, NA nucleus accumbens, PHC parahippocampus. **Attention Networks**: DAN dorsal attention network, INS insula. **Brainstem and Cerebellum**: BS brain stem. **Side-Specific Designations**: LF Left frontal, RF Right frontal LT Left temporal, RT Right temporal, RP right parietal, LP left parietal.

| Study | Sample size | Method/comparison | Findings (tinnitus vs. control) |
| --- | --- | --- | --- |
| (Mohan et al., 2016b) | TN (311) vs. C (256) | **Source Localized**<br>Lagged phase coherence<br>Weighted adjacency matrices<br>Maximum spanning tree<br>Degree<br>Betweenness centrality<br>Eigenvector centrality<br>Closeness centrality | **Common connectivity hubs**<br>Delta: L, SPG, R, S1<br>Theta: L, None. R, IPS<br>Alpha1: L, V1, V2. R, V1, V2<br>Alpha 2: L, V1, V2. R, V1, V2<br>Beta3: L, None. R, dACC, SMA<br>Gamma: Left, SMA, pre-SMA, PCC2, dACC. R, M1, SPS, SMA, pre-SMA, DLPFC, PCC1, PCC2, dACC |
|  |  | Common hubs |  |
| (Mohan et al., 2016a) | TN (311) vs. C (256) | **Source localised**<br>Lagged phase coherence<br>Small world network<br>Clustering coefficient | **Small world network:** TN ↓ delta, alpha1, ↑ alpha2, beta1, gamma.<br>**Node strength:** delta, alpha1 ↑; delta, alpha3, beta1 and gamma ↓<br>**Clustering coefficient:** HC, ↑ delta, alpha1, and in TN ↓ alpha2, gamma.<br>**Local efficiency:** TN ↓ alpha2, beta1, gamma. HC ↑ in delta, alpha1.<br>**Functional distance and path length:** TN ↑ alpha2, beta1 and gamma. HC, delta, theta, alpha1, beta2, and beta3.<br>**Connectivity strength and node pairs:** TN network strength ↓ delta, theta alpha1 and alpha2. ↑ beta1, beta2, beta3 and gamma.<br>**Nodal differences:** TN ↑ delta, theta, alpha2. HC ↑ beta1, beta2 and beta3.<br>**Local efficiency:** TN ↑ beta1, beta2 beta3, gamma. HC ↓ in delta, theta, alpha1 and alpha 2.<br>**Functional distance and path length:** TN ↑ delta, theta, alpha1 alpha2. HC↓ beta1 beta2, beta3, gamma. |
| (Xiong et al., 2023) | TN (57) vs. C (27) | **Source localised**<br>Functional connectivity<br>Lagged Coherence | TN ↑ beta3, R PHC ↔ L PCC (Moderate to severe TN)<br>TN ↑ gamma, R AC ↔ R INS (Moderate to severe TN)<br>TN ↑ gamma, R INS ↔ L PHC & R PCC (correlation to THI score) |
| (De Ridder et al., 2023) | TN (50) vs. C (50) vs. Pain (50) | **Source localised**<br>Lagged phase coherence<br>Granger causality<br>Cross-frequency coupling<br>Phase amplitude coupling | TN ↑ theta pgACC<br>TN ↑ gamma AC ↔ L SMA & M1<br>TN ↑ theta L & R pgACC ↔ mPFC<br>TN ↑ gamma L & R SMA ↔ M1 (compared to pain)<br>TN ↑ theta L & R AC ↔ L & R PHC (compared to pain)<br>TN theta PHC < AC (unidirectional in TN, bidirectional in pain)<br>TN gamma PHC > SOM (bidirectional in pain, unidirectional TN) |
| (Vanneste et al., 2024) | TN on vs. TN off (9) | **Source localised**<br>Lagged phase coherence<br>Granger causality<br>Cross frequency coupling | TN (on) ↓ theta pgACC ↔ AC<br>TN (on) ↑ alpha dACC ↔ pgACC |
| (Mohan et al., 2017) | TN (311) vs. C (256) | **Virtual lesion comparisons**<br>Random node attack<br>Targeted attack (full network)<br>Targeted attack (thresholded networks)<br>Rich club node removal<br>Feeder network node removal<br>Local network node removal | TN ↓ path length<br>TN ↓ global efficiency in low frequency bands<br>TN ↑ short distance in high frequency bands<br>TN ↓ path length, transitivity, global and local efficiency for delta, theta, alpha1, alpha2, beta1, beta2, beta3 and gamma after removal of nodes. |
| (Tsai et al., 2018) | TN (14) vs. C (14) | Dynamic causal modelling | TN ↑ right STG ↔ SPG<br>↑ cuneus/precuneus ↔<br>↑ right cuneus/precuneus ↔ BS all frequency bands except the gamma band.<br>↑ connectivity, superior ↔ transverse STG<br>↑ R side connectivity<br>↑ R cuneus/precuneus compared to L<br>↑ parietal ↔ PFC ↔ NA isthmus of CCG, thalamus, and BS, associated with the perception, emotion, and attention of tinnitus.<br>TN ↑ extrinsic connectivity to the visual and somatomotor systems |
| (Schlee et al., 2009) | TN (21) vs. C (20) | Global phase locking<br>Alpha and gamma coupling | **Long range couplings:**<br>↓ in alpha<br>↑ alpha gamma coupling<br>↓ alpha connectivity LF ↔ ACC, RP, LT<br>RF ↔ LT, LP, PCC, RP, RT<br>ACC ↔ LT, RT, LF<br>LT ↔ LF, RF, ACC, RT, RP<br>LP ↔ RP, RF<br>PCC ↔ RP, RF<br>RP ↔ ACC, LT, LP, PCC, RT<br>RT ↔ RP, LT, ACC, RF<br>↑ gamma LF ↔ RF, RP, PCC, LP<br>RF ↔ LF, LT<br>LT ↔ RF, ACC, RT<br>ACC ↔ LT<br>LP ↔ LF, RT<br>PCC ↔ LF<br>RP ↔ RT, LF |
| (Lan et al., 2021) | Acute TN (< 1 month) (24)<br>Chronic TN (> 6 months), (23) vs. C (32) | Functional connectivity | TN ↑ functional (phase lagged) PHC ↔ PCC PHC ↔ precuneus in chronic but not in acute TN compared to HC or chronic TN. |
| (Zobay et al., 2015) | TN (28) vs. HC (19) | Functional connectivity (Partial directed coherence, imaginary coherence) | TN ↑ effective connectivity AC alpha and beta inflow to the AC from the global network. |
| (Li, et al., 2019) | TN (idiopathic sensorineural hearing loss), (25) vs. C (27) | Functional connectivity, linear lagged connectivity, lagged coherence. | TN ↓ FC in CCG ↔ R PFC<br>↓ gamma FC CCG ↔ FMC<br>↓ supramarginal gyrus ↔ AC<br>↓ DAN ↔ AC |
| (Vanneste, Focquaert, et al., 2011) | Unilateral left-side narrow band TN (18) vs. C (12) | Functional connectivity, multiple voxel-by-voxel t-statistics, with split half reliability analysis. | TN ↓ theta alpha2 gamma,<br>↑ theta, left PHC ↔ right PHC<br>↑ alpha2, L HIP ↔ L AC ↔ R AC ↔ R HIP<br>TN ↑ gamma, L PHC area ↔ L & R AC |
| (Li et al., 2022) | TN (31) vs. C (34) | Functional global connectivity via amplitude envelope correlation | Global connectivity<br>TN ↑ alpha L FP<br>TN ↑ delta LF, L STG, LT, RF, RT, RP |
| (Vanneste & De Ridder, 2015) | Chronic TN (55) vs. C (55) | Seed based lagged phase synchronization | ↑ lagged phase synchronization for alpha 1 PCC ↔ PHC<br>↑ beta3 R insula, PCC ↔ L & R PHC<br>↓ alpha1 L secondary AC ↔ L IFG frontal gyrus ↔ premotor cortex<br>↓ beta3 lagged phase synchronization between left AC ↔ dACC |
| (Pattyn et al., 2018) | TN (with panic disorder), (16) vs. TN (without panic disorder), (16) vs. C (16) | Lagged phase and connectivity analysis. | TN + panic disorder ↓ alpha1 connectivity<br>dACC ↔ insula, amygdala ↔ sgACC in. |
| (Mohagheghian et al., 2019) | TN (8) vs. C (8) | Weighted Phase Lag Index<br>Support vector machine learning classification<br>Network analysis | TN ↑ delta, theta and beta1 connectivity<br>HC ↑ average node strength in alpha2, beta2 and beta3 |
| (Ahn et al., 2017) | TN (16) vs. C (16) | Granger causality<br>Cross frequency coupling<br>Phase amplitude coupling | HC ↑ phase amplitude coupling<br>Frontocentral-phase/Central-amplitude coupling<br>Central-phase/Frontocentral-amplitude coupling<br>Granger causal connectivity was centred on the ACC and DLPFC in all frequency bands.<br>TN ↑ connectivity delta, theta in frontal ↔ AC |
| (Vanneste et al., 2019) | TN (40) vs. C (20) | Lagged phase coherence<br>Granger causality | TN ↑ phase coherence and the connectivity between the PHC ↔ AC in theta and alpha1.<br>TN ↓ phase coherence pregenual anterior cingulate ↔ left HIP<br>Loudness correlated with theta connectivity pgACC ↔ left PHC |
| (Mohan, Davidson, et al., 2020) | TN (311) vs. C (256) | Granger causality<br>Network parameters<br>Functional distance and characteristic path length<br>Clustering coefficient/transitivity<br>Inter/intramodular connectivity<br>Intramodular connectivity<br>Intermodular connectivity | Connectivity strength: TN ↓ alpha1, alpha2<br>Path length: TN ↑ in alpha1, alpha2<br>Transitivity: TN ↓ alpha1, alpha3<br>TN ↓ peripheral nodes, L lateralised connector nodes AC ↔ anterior TN, M1 ↔ ACC<br>TN ↑ path length alpha 1<br>TN ↑ in path length from hubs to left STG ↔ left secondary AC ↔ right rostral ACC<br>↑ path length ↔ right PCC ↔ right rostral ACC all other hubs.<br>↑ alpha2 path length ↔ RT/RP ↔ IFG ↔ left subgenual ACC.<br>Hubs in the tinnitus network;<br>Alpha1<br>Left = ITG, STG, AC<br>Right = PCC, sgACC, dACC<br>Alpha2<br>Left = S1, SMA, Insula, ITG, STG, dACC, sgACC, HIP, AC<br>Right = PreSMA, sgACC, TP, IFG, VLPFC<br>Strength of connections<br>↑ Alpha1<br>Right ACC ↔ Right sgACC<br>↑ Alpha2<br>Right VLPFC ↔ Right IFG<br>Left HIP ↔ Left sgACC<br>Right IFG ↔ Right TP<br>Right IFG ↔ Right VLPFC<br>Left SOM ↔ Left STG<br>Left SOM ↔ Left Insula<br>Left HIP ↔ Right sgACC<br>Left PHC ↔ Left sgACC |
|  |  |  | Left SOM <> Left AC<br>Left ACC <> Right VLPFC<br>Left PHC <> Left HIP<br>Left PHC <> left HIP<br>Right ACC <> right TP |

**Table 3.** Machine learning based studies classification methods and results.

| Study | Number of Participants | Methodology | Features/Technique | Outcome |
| --- | --- | --- | --- | --- |
| **Spectral Frequency Power Based Studies** |  |  |  |  |
| (Wang et al., 2017) | 15 with tinnitus, 14 controls. | Spectral power, support vector machine and multi-view fusion. | Multi-view-fusion feature classification, 6-view-fusion without Alpha 2 band. | Accuracy: 0.828, Precision: 0.811, Recall: 0.857, F1: 0.833 |
| (Vanneste, Song, et al., 2018) | 296 (M), 245 (F), 153 with tinnitus, 264 controls, 78 with pain, 31 with Parkinsons, 15 with major depression. | Current density of all frequency bands at regions of interest. Support vector machine learning classification. | Contributors to the model:<br>Theta, beta, and gamma power at the dorsal anterior cingulate cortex, posterior cingulate cortex, auditory cortex, and the gamma-frequency band for the subgenual anterior cingulate cortex and parahippocampus. | Accuracy: 87.7%, TPR: 0.82, FPR: 0.08 |
| (Mohsen et al., 2023) | 20 (M), 9 (F), with tinnitus | Logistic regression supervised machine learning. | Source reconstructed frequency bands (Delta, Alpha), mean total of electrodes | Accuracy: 0.80, Sensitivity: 0.86, Specificity: 0.69 |
| **Connectivity Based Studies and Network Analysis** |  |  |  |  |
| (Schlee et al., 2009) | 12 with tinnitus, 20 controls. | Phase locking and inter-areal coupling | Alpha and gamma coupling (networks) | Discrimination accuracy: 83% |
| (Li et al., 2022) | 31 with tinnitus (20 (M)), 34 controls (13 (M)) | Support vector machine, and multi-layer perceptron network. | Phase locking values, Pearson correlation coefficients | Accuracy: 99.4% |
| (Wang et al., 2023) | 187 tinnitus patients, and 80 controls | Multi-Band EEG Contrastive Representational Learning, best compared to other models. | Alpha1, Beta1, Beta2, Beta3 in default mode network. High value electrodes are (Fp1 and Fz), parietal (Pz), central cranial (Cz), temporal (T5 and T3) and occipital region (O1) in each frequency. | Precision: 94.4, Recall: 95.0, F1: 94.7 |
| **Other Features** |  |  |  |  |
| (Allgaier et al., 2021a) | 29 with tinnitus and 13 controls. | Neural Networks | 1-second noise-reduced, down-sampled raw data epochs | Accuracy: 75.6% |
| (Emami & Bayrak, 2017) | 6 with tinnitus and 6 controls | Comparing machine learning models in the time domain after noise reduction. | K Nearest Neighbour with K = 3 and distance metric: Euclidean, Distance weight: square inverse | Accuracy: 95.5%. Best epoch is 8 seconds, inverse weighting distance is the best distance function. |
| (Jianbiao et al., 2023) | 10 with tinnitus and 10 controls | Support vector machine | Frequency band entropy values (increased values in tinnitus) | Accuracy: 91% |
| (Piarulli et al., 2023) | 129 tinnitus patients and 142 controls | Support vector machine | Source reconstructed: absolute power spectral densities, relative power spectral density, complexity, inward flow, outward flow features. | Tinnitus accuracy: 94-97%, Distress accuracy: 84-89% |
| (Ahmed et al., 2024) | 51 deaf patients, 54 with tinnitus and 42 controls | Principle component analysis, feature reduction, evaluated with ensemble learning techniques; Random Forest, Extra Trees, Gradient Boosting, and CatBoost | Frequency band power data set. Extra Trees, and CatBoost performed best. | Precision and recall: ~89% and F1-score: 89.31 |

**Table 4.**
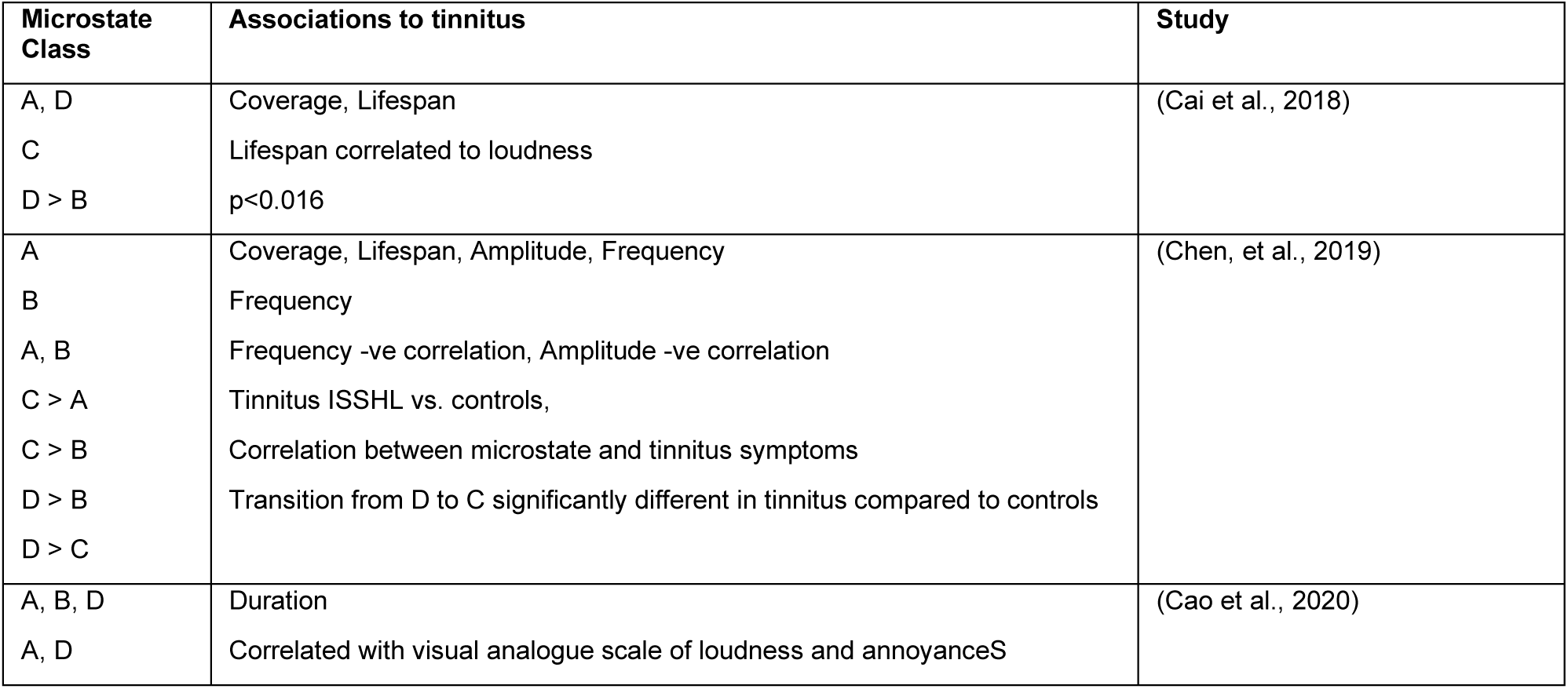
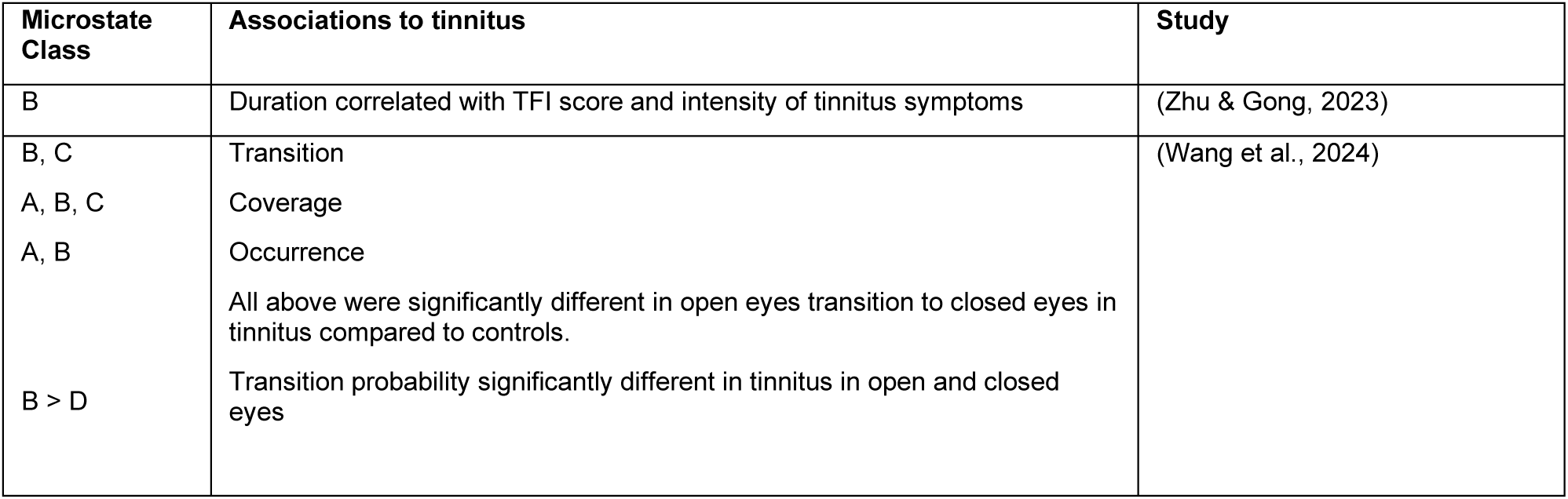
Microstate analysis of tinnitus. A is associated with the regions right-frontal left posterior, B is associated with left-frontal right posterior, C is associated with anterior-posterior, and D associated with fronto-central extreme. > denotes transition between states being significantly different.

### 3.1 Spectral Band Power Studies

The most common analysis evaluated differences in EEG spectral power between tinnitus and controls, from scalp based electrodes, either at the cortical level (Klimesch, 2018), or in inferred subcortical sources reconstructed from the scalp electrode signals (Pascual-Marqui, 2002; Pascual-Marqui et al., 1994). The power spectra are commonly divided into bands named delta (∼1–4 Hz), theta (∼4–8 Hz), alpha (∼8–13 Hz), beta (∼13–30 Hz), and gamma (∼30–45 Hz).

Numerous studies further subdivide these bands (e.g. low and high alpha, beta or gamma). For this scoping review, we merged subdivisions into the canonical band names as listed above (Figure 3, Table 2 & Table 3). Despite differences in spatial accuracy, for the purposes of evaluating potential objective markers of tinnitus, this review combines frequency-based results using sensor locations and source reconstruction. The data for Figure 4 is in Appendix 2.

**Figure 3.**
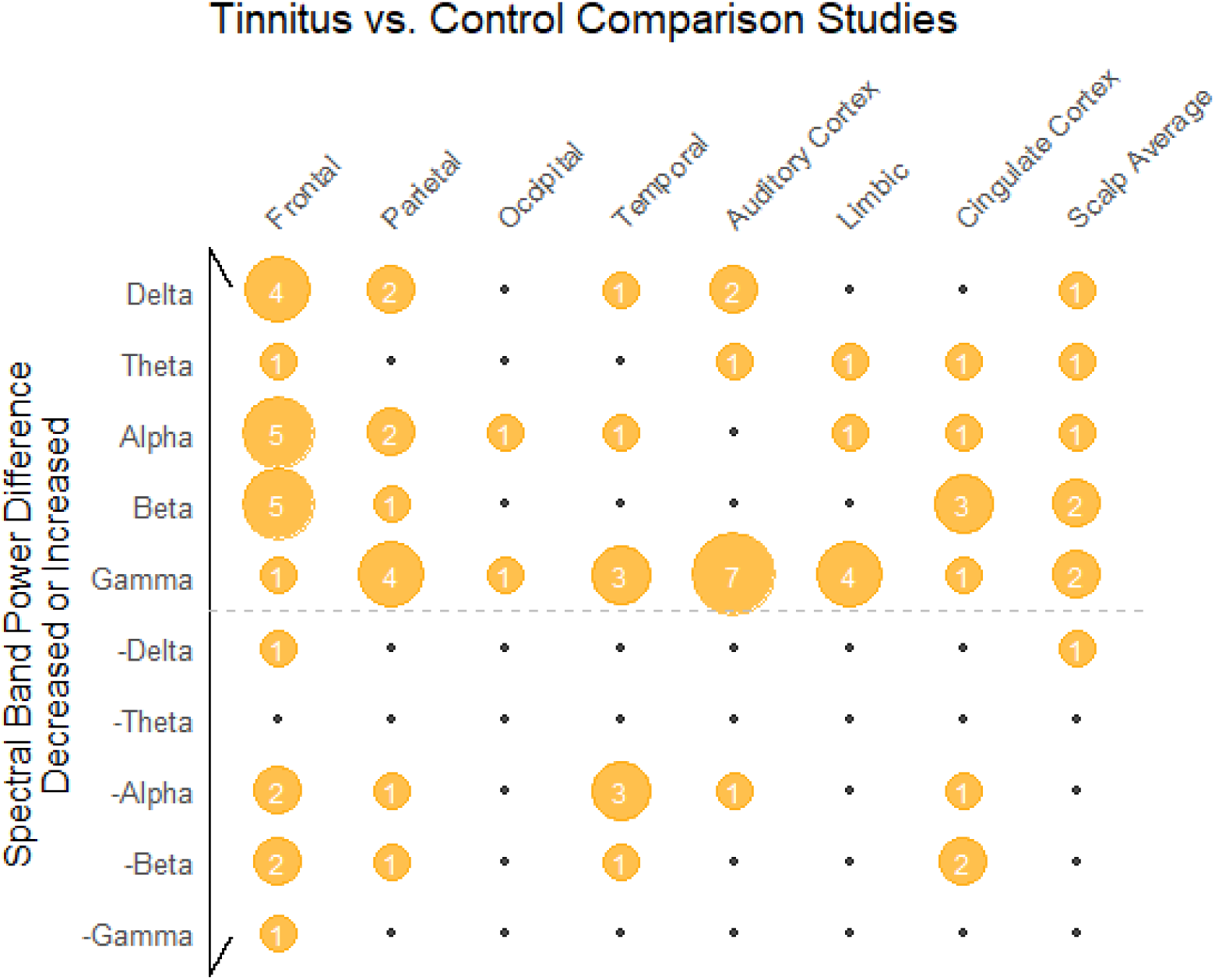
The numbers indicate the count of studies reporting difference in spectral power by band and brain region in tinnitus vs. control group. Only studies reporting differences associated with tinnitus improvements of at least 50% were included. All locations are estimated based on sensor location or source location as appropriate. Increases in band power are shown above the dotted line, and decreases in band power are below the dotted line. Only studies that reported statistically significant findings are counted in this figure (quality of the statistics was not evaluated). Most studies reported multiple regions and frequency power differences.

**Figure 4.**
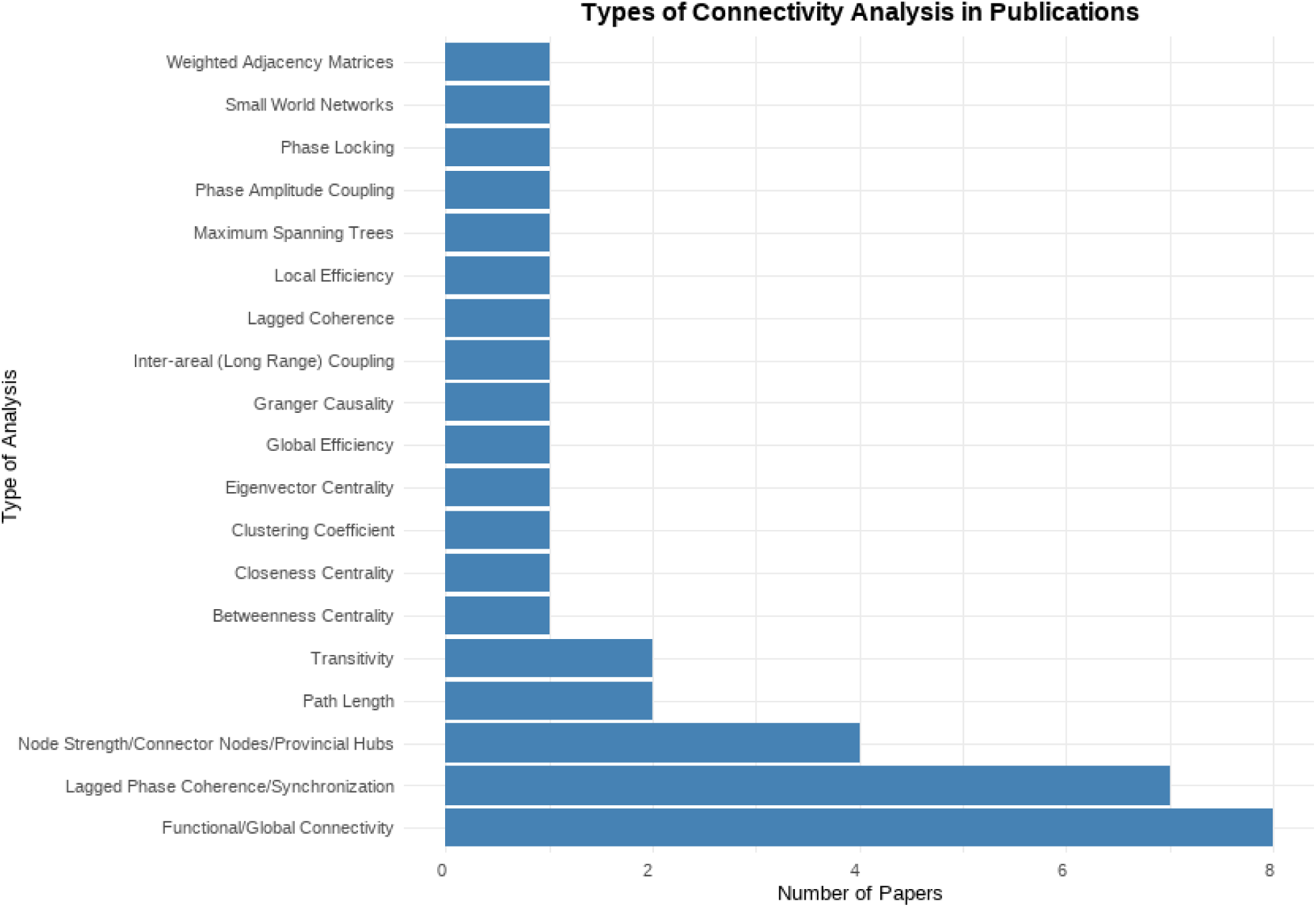
Count of types of connectivity analysis, most papers used multiple approaches (Total = 20).

### 3.3 Connectivity Group Comparison Studies

Network connectivity features are network characteristics such as node strength, hubs, edge characteristics and information exchange, as well as a range of other related features (Chiarion et al., 2023).

The results from studies comparing tinnitus to controls are in Table 2. Functional and global connectivity relates to analysis of parts of the brain that are functionally connected and is evaluated by measuring the similarity of activity in pairs of regions. Nine studies compared network and connectivity across a range of features of tinnitus (high and low distress, age of onset, hearing loss, laterality and tinnitus frequency) are reported in Appendix 1.

### 3.4 Feature-based machine learning

Studies that used either a combination of features for classification, or unique features not mentioned in the sections above, or did not identify the direction of difference, or if they compared multiple machine learning techniques as their focus are summarised in Table 3.

### 3.5 Information Theory Measures

Entropy can be a measure of how much information a dynamic system is carrying in its signals, and coding efficiency, particularly of sensory neurons. Sadeghijam et al, (2021) found increased entropy in a tinnitus subject group compared to controls across all regions of interest, particularly in the high alpha frequency band and beta in the right auditory, right frontal and central regions. In another study using machine learning, delta, alpha1 and beta1 band sample entropy values were greater in tinnitus compared to controls, in the parietal, central and left prefrontal regions (Jianbiao et al., 2023), Table 3.

### 3.6 Microstates

The brain is a dynamic system, and a microstates analysis attempts to capture the features of large scale network dynamics in the brain (Khanna et al., 2015). The studies in Table 4 indicate that there are differences in cortical activity states and state changes between tinnitus and controls. Microstates are difficult to assign directly to a functional component or network, so the conclusions beyond differences in neural dynamic states are limited.

## 4. Discussion

This review evaluates resting-state EEG features as potential objective measures of chronic subjective tinnitus. The findings highlight a complex interplay of neural dynamics, with differences in spectral band power, connectivity, and network characteristics across auditory and non-auditory brain regions in people with tinnitus compared to controls. These observations underscore the multidimensional nature of tinnitus and the challenges in identifying reliable objective measures.

This section discusses key findings, the role of machine learning in advancing tinnitus research, and implications for future studies and clinical applications.

### 4.1 Neural Dynamics and EEG Features in Tinnitus

Resting-state EEG studies comparing individuals with tinnitus to controls reveal widespread differences in scalp electrode source derived neural activity, particularly in the anterior cingulate cortex, prefrontal cortex, auditory cortex, and limbic system—regions associated with auditory processing, attention, memory, and emotion (Moring et al., 2022; Ueyama et al., 2013). Spectral band power analyses (Figure 3) indicate consistent increases in auditory gamma band power, alongside variable differences in frontal alpha, beta, and delta bands across these regions (Figure 2). These findings align with recent MEG reviews reporting enhanced alpha and beta connectivity between frontal and auditory regions in those with tinnitus with and without hearing loss (Reisinger et al., 2023). The variability in spectral power differences is likely influenced by secondary factors such as hearing loss, psychological distress, coping styles, and tinnitus sound frequency (Appendix 1). These factors highlight that tinnitus is not an isolated phenomenon but is embedded within broader neural and psychological dynamics (Husain & Khan, 2023). Source-derived EEG measures further suggest involvement of deeper brain regions, including those linked to memory and emotion, indicating that hierarchical sensory integration networks may contribute to tinnitus perception (Sedley et al., 2016; Vanneste & De Ridder, 2012), however, source estimation of cortical structures is an approximation. These complex neural patterns pose challenges for identifying consistent objective measures but also point to potential therapeutic targets.4.2 Challenges in Standard Methodological Approaches Standard EEG analysis methods, such as spectral band power and connectivity analyses, have been instrumental in identifying neural correlates of tinnitus. However, these approaches face limitations due to the heterogeneity of tinnitus populations and methodological variability across studies. For instance, differences in directing participants to attend to or ignore their tinnitus percept can affect EEG recordings, emphasizing the need for standardized protocols to ensure high-quality data (Adjamian et al., 2016). Additionally, the complexity of matching control groups— given variations in tinnitus symptoms, hearing loss, and comorbidities like depression or pain— complicates the identification of reliable biomarkers. These challenges highlight the limitations of traditional statistical methods, which often rely on predefined assumptions and may struggle to capture the multidimensional nature of tinnitus.

### 4.3 Advantages of Machine Learning Approaches

Machine learning classification techniques offer significant advantages over standard methodological approaches for identifying objective tinnitus measures. Unlike traditional methods, which often focus on isolated EEG features (e.g., spectral power in specific bands), machine learning can integrate multiple features—such as spectral band power, network connectivity, microstates, and entropy—into a single model (Figure 4). This multidimensional approach is better suited to capturing the complex, heterogeneous neural signatures of tinnitus. For example, machine learning studies utilizing support vector machines and neural networks have successfully differentiated tinnitus from healthy controls and related conditions like pain and depression, achieving promising classification accuracies (Table 3).

Machine learning approaches excel in handling large, diverse datasets and identifying patterns that may not be apparent through conventional analyses. By combining spectral and connectivity measures, machine learning models have demonstrated superior performance in distinguishing tinnitus-specific neural patterns (Althnian et al., 2021). Moreover, machine learning’s ability to process high-dimensional data enables the exploration of understudied EEG features, such as microstates and entropy, which may reveal novel insights into tinnitus mechanisms. In contrast to standard methods, which often require manual feature selection and may miss subtle interactions, machine learning algorithms can automatically identify relevant features and their interactions, enhancing predictive power.

Another key advantage of machine learning is its potential for interpretability when paired with explainable AI techniques (Shabestari et al., 2025). Interpretable machine learning models, grounded in theoretical frameworks like the Bayesian Brain model (Hu et al., 2021), can elucidate how specific EEG features contribute to tinnitus classification or subtyping (De Ridder et al., 2023), and predict responses to interventions (Cardon et al., 2022), and progression (Hobeika et al., 2025). This interpretability is critical for clinical translation, enabling researchers to link machine learning findings to underlying tinnitus mechanisms and evaluate and predict the impact on neural measures following interventions. However, challenges remain, as 7 of the 12 machine learning studies reviewed had small sample sizes (n < 35), limiting generalizability. Larger, more diverse datasets are needed to enhance model robustness and clinical applicability.

### 4.4 Future Directions and Clinical Implications

A number of tinnitus models have been proposed, (Biehl et al., 2019; Gerken, 1996; Noreña, 2011; Schlee et al., 2011; Sedley et al., 2016; Vanneste, Song, et al., 2018) and it is important to consider the predictions of these models in the context of differences in EEG; the factors that may impact differences, and the outcome measurement methods. The integration of machine learning with EEG-based tinnitus research holds substantial promise for identifying objective measures and advancing clinical applications (Allgaier et al., 2021b). Focused machine learning assessments of spectral power, connectivity measures, microstates, entropy and other non-oscillatory features (Kowalik & Elbert, 1994) could refine our understanding of tinnitus subtypes and neural mechanisms. Recent studies using conjunction analysis have distinguished tinnitus from pain networks based on Bayesian models, suggesting machine learning could further elucidate these differences (De Ridder et al., 2023). Developing interpretable machine learning models that align with neurobiological models of tinnitus is a critical next step for translating findings into clinical tools for diagnosis, subtyping, and monitoring treatment outcomes.

### 4.5. Limitations

This scoping review focussed on resting-state EEG and comparisons between participants with chronic subjective tinnitus and controls. However, there are limitations that bear consideration.

#### 4.5.1 Quality of Studies

We did not assess in detail the methodological quality of the studies included in this review. A range of methodological differences can alter observed EEG activity (Adjamian et al., 2016). In addition to methodological differences, differences relevant to subtyping such as hearing status (Adjamian et al., 2012), gender, age, distress and depression (Riha et al., 2022) and various comorbidities were also not a focus of this review, but are important for subtype analysis. (Meyer et al., 2017) see (Appendix 1).

#### 4.5.2 Feature Reporting

We sought broad changes in features such as spectral power and spectral density and did not differentiate source reconstructed and sensor-based locations as brain regions. Differences in spectral density may have been absolute or relative changes, corrected or uncorrected (for multiple comparisons) or classifier features.

#### 4.5.3 Type of Studies

Studies that evaluated evoked response evidence were excluded. Neural event response studies are a large area of research, and a large and valuable area for consideration. Event related potential studies may contain candidates for objective measures of chronic subjective tinnitus (Fabrizio-Stover et al., 2024). Intervention studies were not included due to the impact of the intervention on EEG characteristics, and the lack of relationship to tinnitus measures. This is a rich area of research for monitoring changes evoked by experimental interventions.

#### 4.5.4 Exclusion of MEG and EcoG

The intention of focussing on EEG measures with potential direct translation to clinical use therefore excluding MEG and EcoG research, however a recent review (Reisinger et al., 2023) reached similar conclusions to this review in terms of future novel approaches (machine learning), and methodological standardisation (Adjamian et al., 2016).

### 4.6. Future Directions

The complexity of identifying objective measures for chronic subjective tinnitus, complicated by its impact on various neural networks, symptom heterogeneity, and individual variability, poses significant research challenges. To tackle these:

- **Model-Based Hypotheses**: Construct studies around hypotheses informed by theoretical models such as the Bayesian Brain model (De Ridder et al., 2023, 2024; De Ridder & Vanneste, 2021), central gain model (Auerbach et al., 2014; Hutchison et al., 2023; Noreña, 2011; Zeng, 2013), or global brain model (Hazell & Jastreboff, 1990; Schlee et al., 2011; Weisz, Dohrmann, et al., 2007). Refining these models has led to a level of convergence of ideas in sensory precision (Hullfish et al., 2019; Sedley et al., 2016), testing the predictions of these ideas with experimental EEG data is vital for progress in tinnitus research.
- **Study Design**: Prioritize well-powered studies with diverse participant pools, employing standardized methods to simplify comparison across studies (Adjamian et al., 2016). The inherent subjectivity of tinnitus may bias participants to be those who find it subjectively worse but may be objectively the same. Designs that include comparison of risk factors that increase severity with EEG features will potentially enable early intervention and severity tracking longitudinally (Hobeika et al., 2025).
- **Statistical Robustness**: Implement statistical methods like linear mixed effects models to manage variance and individual differences (Cederroth et al., 2019; Riha et al., 2020), to account for heterogeneity in tinnitus presentation.
- **Machine Learning**: Leverage the potential of machine learning, particularly with sophisticated algorithms like support vector machines and neural networks, to identify, classify, and track differences in EEG features related to chronic subjective tinnitus symptoms such as loudness, annoyance, and distress (Allgaier et al., 2021; Emami & Bayrak, 2017; Mohagheghian et al., 2019; Vanneste, Song, et al., 2018; Wang et al., 2017). Utilising emerging explainable AI techniques that combine risk factors and machine learning classification techniques (Shabestari et al., 2025) can assist in the classification of subtypes, and pinpointing opportunities for effective clinical intervention.
- **Sharing Datasets**: Sharing high-quality, diverse datasets is crucial for further validation of these tools. If future research can release anonymised datasets in the open-source domain, machine learning specialists can explore various features and approaches.
- **Future Systematic Review Potential**: Heterogeneity in tinnitus research methods and subtypes remains a distinct issue restricting progress to useful clinical measures of tinnitus. Tinnitus EEG researchers would benefit from future systematic reviews that 1) Target evidence from tinnitus models 2) Delineate evidence for subtypes in EEG and 3) Based on models, evaluate EEG features to specific chronic subjective tinnitus measures.

## 5. Conclusion

In summary, this review highlights the impact of chronic subjective tinnitus on EEG metrics. Spectral band power, connectivity, and network characteristics emerged as key areas for assessing tinnitus symptoms, separating subtypes, monitoring symptom changes, and aiding in the development of targeted therapeutic strategies. Systematic testing based on theoretical models, using diverse datasets, robust statistical methods, and interpretable machine learning algorithms, is essential for identifying clinically-useful objective measures of tinnitus.

## Data Availability

All data produced in the present work are contained in the manuscript

## Key Terms and acronyms

EEG: electroencephalography
DLPFC: dorsolateral prefrontal cortex
fMRI: functional magnetic resonance imaging
TFI: Tinnitus Functional Index

## APPENDIX

**Appendix 1:** Connectivity differences when comparing between various tinnitus groups or with controls.

2 Appendix 1: Connectivity differences when comparing between various tinnitus groups or with controls.
| Paper | Comparison | Summary of network differences |
| --- | --- | --- |
| (Mohsen et al., 2019) | Chronic Tinnitus split into low vs. high (>56 tinnitus handicap index score) | Elevated functional coherence connectivity in the high tinnitus handicap index group between the secondary auditory cortex and the frontal areas, and the parahippocampus and centrofrontal regions. |
| (Song et al., 2013) | Early onset vs. late onset tinnitus and control groups | Increased localized activity in distress networks and default mode networks, and intrinsic alertness networks. Differences had similar features to pathologies to pain and depression. |
| (Demopoulos et al., 2020) | Tinnitus with and without hearing loss and control groups | Global functional connectivity changes associated with tinnitus. Beta connectivity may be associated with non-auditory symptoms and different between those with and without hearing loss. |
| (Song et al., 2015) | Onset age. Early onset, compared to late onset. High distress compared to low distress. | Increased functional connectivity for the late onset, primarily in the anterior cingulate cortex and parahippocampus. Early-onset tinnitus with high distress was associated with the orbitofrontal cortex and dorsolateral prefrontal cortex connectivity. similarities of the anterior cingulate cortex networks to depression networks. |
| (Vanneste, Plazier, et al., 2011) | Left sided vs right sided tinnitus compared to controls | No differences in auditory cortex connectivity. |
| (Vanneste et al., 2014) | Coping style comparison, adaptive vs maladaptive coping styles. | Increased activity in resting-state connectivity in the default resting-state networks in those with a maladaptive coping style. Alpha band hyperconnectivity in the default network in those with maladaptive coping style. |
| (Vanneste & De Ridder, 2015) | Loudness, grades of tinnitus related distress. | Loudness, distress and increase in grade of tinnitus related distress result in the activation of different network connections at different frequency bands. |
| (Vanneste & De Ridder, 2016) | Tinnitus with and without hearing loss | Two different tinnitus generating network mechanisms depending on the amount of hearing loss, with tinnitus being associated with parahippocampal involvement and the auditory cortex implicated in tinnitus with no hearing loss. Increased functional and effective connectivity between the auditory cortex and the parahippocampus. |
| (Zhang et al., 2020) | Low and high frequency tinnitus | Higher gamma linear connectivity between the right primary and secondary auditory cortex and the right angular gyrus in the high frequency group. |
| (Vanneste et al., 2024) | Tinnitus on vs. tinnitus off | Increased theta-gamma coupling between the pregenual anterior cingulate cortex and the auditory cortex. Alpha effective connectivity increased from the dorsal anterior cingulate cortex in tinnitus on. |

**Appendix 2:**
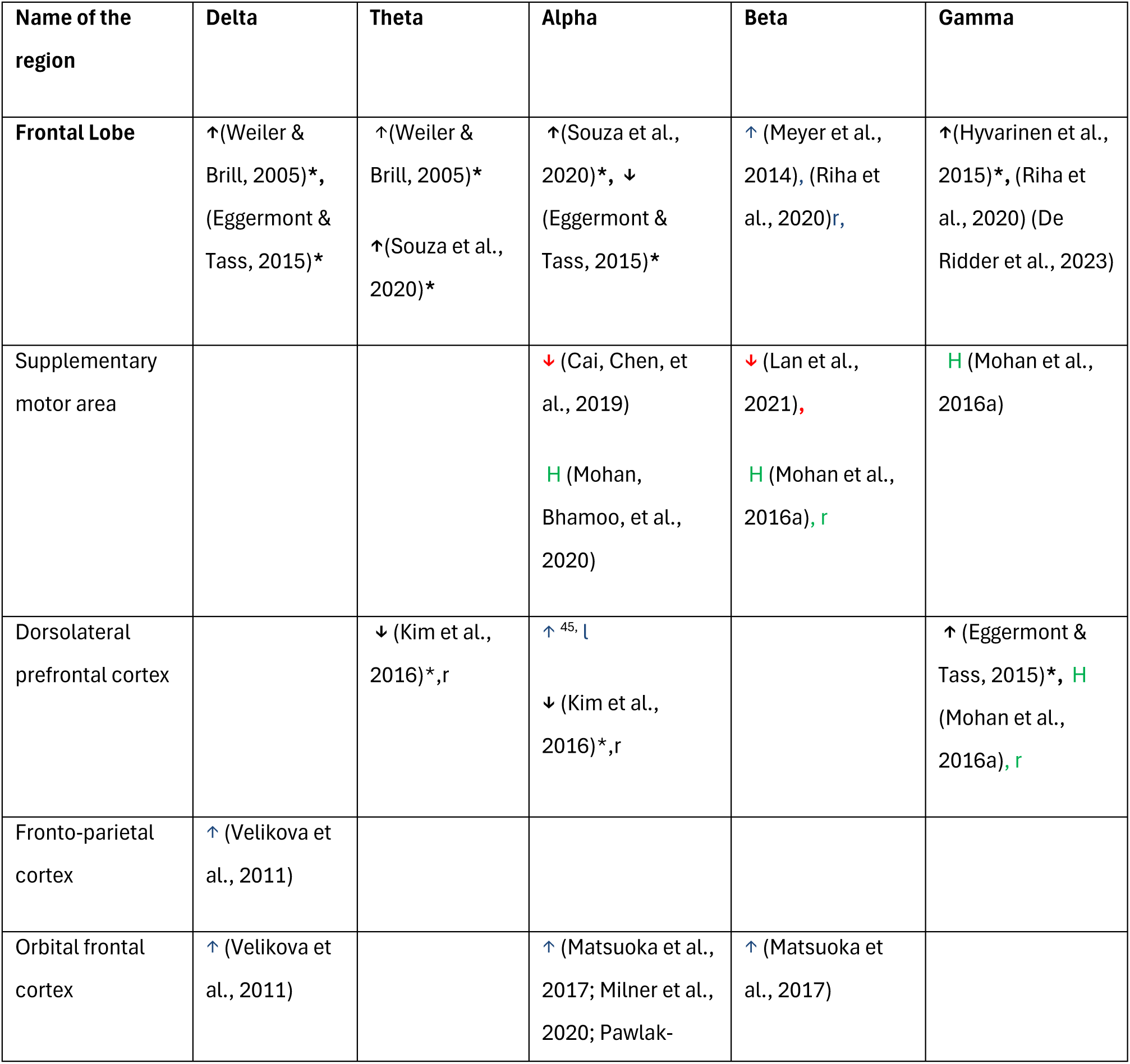
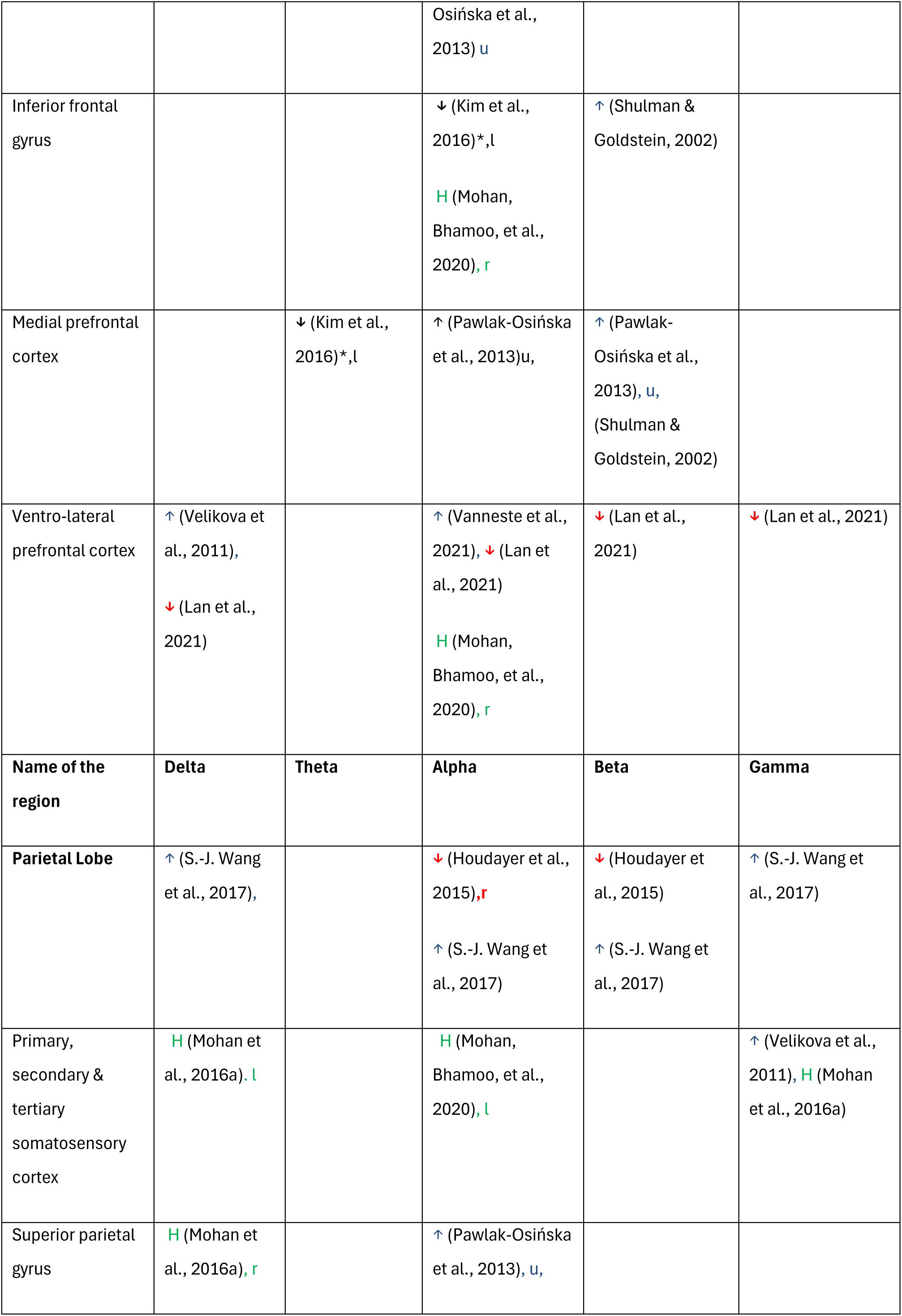
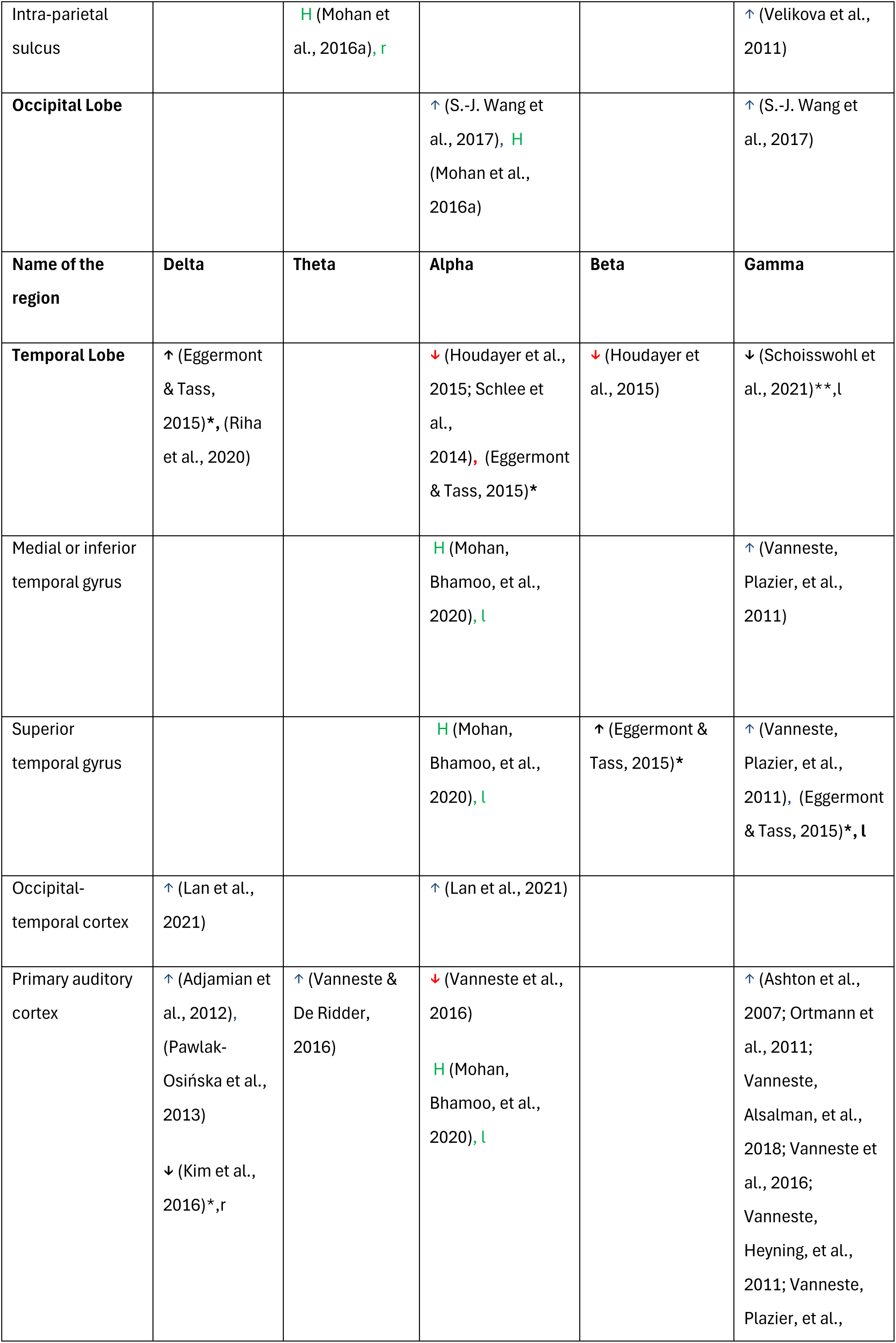
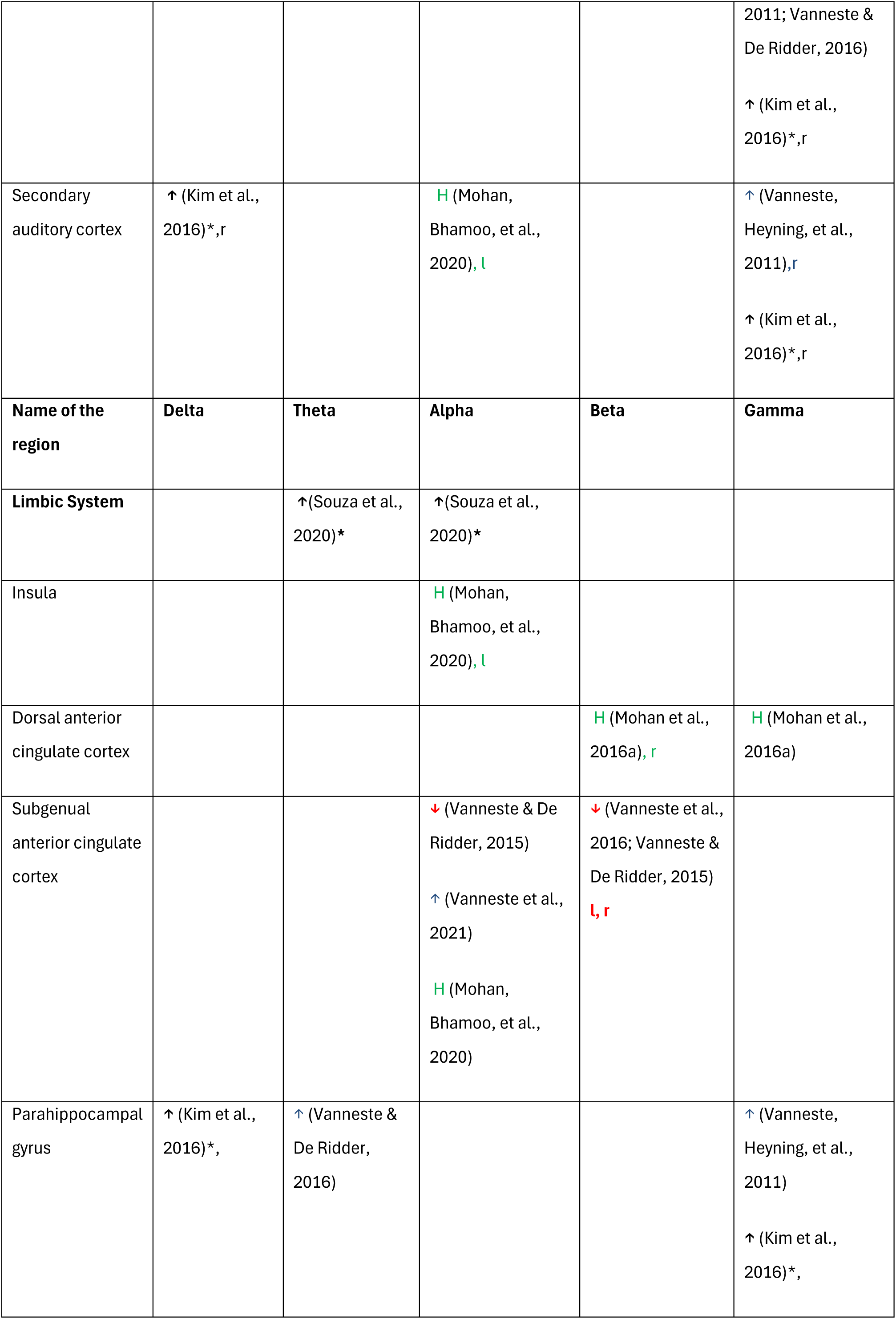
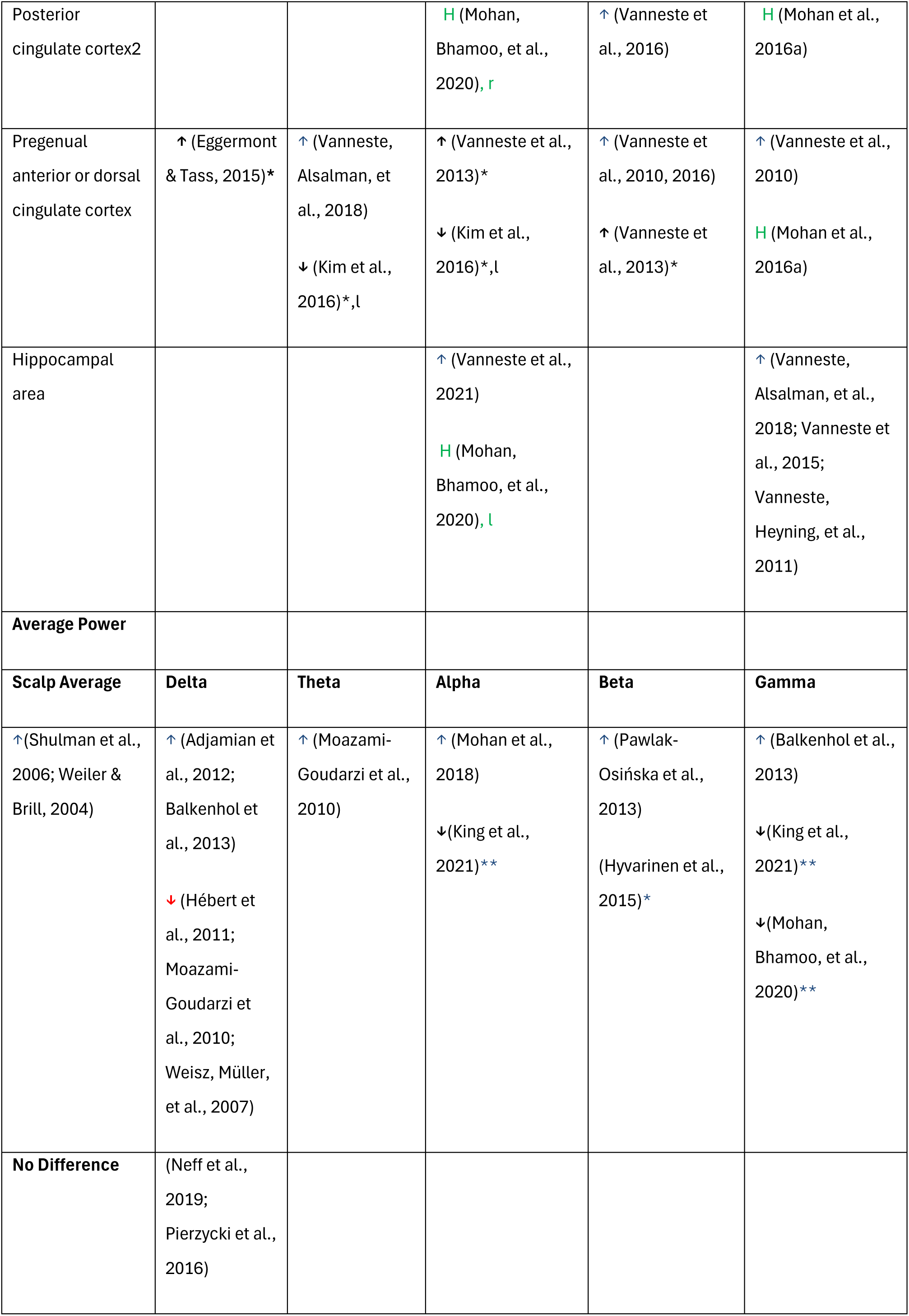
Details of the spectral power in tinnitus compared to controls by region and connectivity hubs. Note reported electrodes were allocated to the nearest region. Both source analysis, and electrode locations are included. ↑=higher power in tinnitus, ↓=lower power in tinnitus, r=right side, l=left side, u=uncorrected. * = Intervention associated with tinnitus change (note only studies that reported more than 50% improvement in tinnitus are included), higher power or lower power are in comparison with after treatment. **=Intervention led to increased alpha + gamma in responders (↓=lower power in tinnitus compared to non-responders). Red and blue are resting-state EEG frequency studies, black are intervention studies. H = connectivity hubs. Green = hubs, red and blue = frequency studies, **black** = intervention studies.

